# The global return-on-investment of COVID-19 vaccines in the first year of the vaccination programme

**DOI:** 10.1101/2025.09.02.25334932

**Authors:** Hallie Benjamin, Gregory Barnsley, Oliver J Watson, Mark Jit

## Abstract

COVID-19 vaccines played a critical role in reducing global health burden during the COVID-19 pandemic, but their rapid development required extraordinary effort. This study aims to evaluate their global return-on-investment (ROI) during the first year of the vaccination programme (8 December 2020-8 December 2021) to provide insights into the economic benefits relative to the costs of vaccine development, manufacturing, and delivery. Public health outcomes of COVID-19 vaccination, including life years and QALYs gained from averted death, infections, and hospitalisations, were estimated. Health benefits were converted to economic benefits using welfarist and extra-welfarist approaches. These benefits were compared to the costs of vaccine development, manufacturing, and delivery to estimate the ROI under both approaches. Probabilistic sensitivity analyses were conducted to assess the impact of parameter uncertainty on ROI estimates. Vaccine development and delivery cost $79.4 billion, but the health and economic benefits of vaccination were valued between $4.83 trillion–$37.8 trillion. High-income countries saw the greatest health benefits per-person vaccinated, and low-income countries gained the greatest non-health benefits per-person vaccinated, when expressed as a proportion of GDP per capita. This estimated ROIs of $59.8–$475 per dollar invested. ROIs ranged between $62.2–$442 across all sensitivity analyses for both approaches. Rapid COVID-19 vaccine development and delivery were an excellent investment. Vaccine development and equitable vaccine distribution globally should be prioritised before and during global health crises.

## Background

After SARS-CoV-2 was identified as the novel coronavirus responsible for COVID-19 in 2020, an extraordinary global effort was launched by governments, pharmaceutical companies and non-profit organisations to rapidly develop vaccines. Remarkably, the first COVID-19 vaccines received emergency use authorization within 12 months of viral identification, a process that typically takes a decade or more. These vaccines proved to be highly effective in preventing severe illness and death, with a modelling study finding that they averted between 14-20 million COVID-19-related deaths in the first year of use.^1^ However, despite their clear health impact, the economic value of these vaccines – particularly in relation to the massive investments in their rapid development – remains unexamined on a global level.

Although numerous economic evaluations of COVID-19 vaccines have informed vaccine procurement and deployment at the country-level, none have assessed the value for money of vaccine development itself, rather than just vaccine procurement. A systematic review of economic evaluations for COVID-19 vaccines conducted in 2023^2^ identified 25 full economic evaluation studies, including cost-minimization, cost-benefit, cost-effectiveness, and cost-utility analyses. However, they all compared country-level benefits to the cost of the national programme (rather than the cost of vaccine development).^2^ While a recent study looked at COVID-19 vaccination’s impact on global gross domestic product based on regression analysis, it did not look at the return-on-investment (ROI) of vaccine development itself.^3^

ROI analyses evaluate the economic benefits of an intervention relative to its costs, often using a cost-benefit approach that monetizes health and non-health impacts.^4^ Despite the complexity of conducting ROI analyses for vaccine impact, including selecting the appropriate methodology, ROI analyses are vital for demonstrating the health and economic value of vaccines and building strong investment arguments.^5^ The ROI of COVID-19 vaccine development is an important unanswered question because the unprecedented effort in developing, manufacturing, and distributing COVID-19 vaccines to achieve high coverage is increasingly being questioned. A Pew Research survey found that a substantial minority of U.S. respondents were skeptical about the health benefits of COVID-19 vaccines, with notable levels of distrust in mRNA vaccines also observed in other countries.^6,7^ Recently, there have been promising steps toward greater equity in pandemic vaccine distribution, such as the signing of the World Health Organization’s Pandemic Treaty.^8^ However, some funding streams for pandemic vaccine research have also been cut.^9^ In the event of future pandemics, a clearer understanding of the ROI from COVID-19 vaccines could play a critical role in guiding decisions around comparable investments in rapid vaccine development. Furthermore, COVID-19 vaccine benefits are unequally distributed across countries due to access inequalities, and the economic implications of this inequity have not been fully enumerated.

To address these evidence gaps, we aimed to evaluate the global ROI for COVID-19 vaccines over the first year of their use (8 December 2020–8 December 2021), when the vaccines had the greatest impact due to limited pre-existing immunity, by comparing the vaccines’ benefits to their development and distribution costs. We also aimed to estimate the relative contribution of the wider societal benefits of COVID-19 vaccination, such as reducing morbidity, mortality, and productivity, to the overall ROI, and to characterise the differences in the economic and health benefits generated between World Bank income groups.

## Methods

### Overview

The ROI of COVID-19 vaccination over the first year of the vaccination programme globally was determined by monetising its public health outcomes and then comparing them to the investments made into COVID-19 vaccine development and delivery using two standard approaches: a welfarist and an extra-welfarist approach.^10^ The welfarist approach uses the value of statistical life (VSL), which captures individuals’ valuations and preferences for reducing their own mortality risk, whereas the extra-welfarist approach broadens the evaluation to capture the inherent value society places on saving a life and being in good health. For this approach we also include the value of improved economic productivity and reduced healthcare costs. Productivity and healthcare costs are not included in the welfarist approach as these costs are assumed to be factored into individuals’ valuations of their preferences for reductions in mortality risk.

To accurately assess the impact of COVID-19 vaccination on mortality and morbidity, we used the results of a global transmission model-based analysis of COVID-19 vaccination by Watson et al.^1^. This analysis provided estimates of the number of infections, hospitalisations, and deaths averted due to COVID-19 vaccination efforts worldwide for seventeen discrete age groups (0-5, 5-10, …, 75-80, 80+) (Table 1). Other data required for our analysis were collected from online secondary sources, including the World Bank, World Health Organization (WHO), and published papers (Table 1). Our analysis includes 173 countries (i.e. the 180 countries analysed by Watson et al, except for countries lacking World Bank economic indicators (Cuba, Eritrea, North Korea, South Sudan, Venezuela, and Yemen) or WHO life expectancy data (Aruba)).

**Table 1.** Input parameters used in the economic analysis.

| Parameter | Sources | Notes and assumptions |
| --- | --- | --- |
| **Health benefits** |  |  |
| COVID-19 vaccine impact on public health outcomes, including averted deaths, infections and hospitalisations | Watson et al.<sup>1</sup> | Available for discrete age groups (0-5, 5-10, ..., 75-80, 80+) for 173 countries. |
| Cohort life expectancy | WHO Global Health Observatory indicator views<sup>30</sup> | Dependent on country and age group. |
| Discount rate | WHO recommendation<sup>31</sup> | Assumed 3% and 0%. A discount rate is applied to determine the current value of costs and benefits that occur in the future, which enables the comparison of costs and benefits that occur at different times. |
| Quality-adjusted life years (QALYs) lost due to COVID-19 infections, hospitalisations and deaths | Basu et al.<sup>32</sup> | QALY losses were estimated for COVID-19 based on patients experiencing severe influenzas or H1N1. |
| Infection and hospitalisation durations | Watson et al.<sup>1</sup> | To be consistent with original modelling study, we assumed that infections had a duration of 5 days, and hospitalisations had a duration of 12 days. |
| **Monetising health benefits** |  |  |
| Reference USA value of statistical life (VSL) | United States Department of Agriculture Economic Research Service<sup>33</sup> | Estimate of VSL for 2020 used for analysis |
| Gross national income per capita | World Bank<sup>34</sup> | Atlas method |
| Income elasticity | Masterman et al.<sup>35</sup> | Assumed value of 1 |
| Gross domestic product (GDP) | World Bank<sup>36</sup> | Estimate of GDP for 2020 used for analysis |
| Percentages of GDP for willingness-to-pay thresholds | Pinchon-Riviere et al.<sup>37</sup> | Represents health opportunity costs in each country, i.e. how much health spending results in gaining a year of life in perfect health |
| **Productivity costs** |  |  |
| Gross domestic product per capita (GDPpc) | World Bank Data<sup>38</sup> | GDPpc divided by 365.25 days was used as a proxy for average daily income.<sup>39</sup> Estimate of GDPpc for 2020 used for analysis. |
| Friction periods | Low- and middle-income countries (LMICs): Krol and Brouwer<sup>39</sup> | A friction period of 3 months was used for LMICs as recommended by Krol and Brouwer (2014), due to a lack of reliable friction period estimates. |
|  | High-income countries: Hanly et al.<sup>40</sup> | Given that estimated friction periods were provided for only Organisation for Economic Co-operation and Development countries, to improve consistency with LMIC data, the European average friction period value of 60.6 days was used. |
| Healthcare costs | LMICs: Torres-Rueda<sup>41</sup> 2025-9-2 10:32:00 AM | Used the “Hospital-based (severe cases) per day of hospitalisation (severe case)” values. |
|  | HICs: WHO-CHOICE<sup>42</sup> | Used the “Cost per inpatient bed day by hospital level (without drugs)” values for primary hospitals. |
| GDP deflator indices | World Bank<sup>43</sup> | Used for 2010, 2019, 2021, and 2022. |
| **Vaccine development and delivery costs** |  |  |
| Vaccine development costs (including Advanced Purchasing Agreements, corporate investments, and manufacturing costs) | Wouters et al.<sup>14</sup><br>Florio et al.<sup>15</sup><br>Publicly available data<sup>16</sup> | Development costs include investments made by public and private sectors, philanthropic and other funders, either directly to organisations and companies conducting research, development and manufacturing, to intermediaries or through Advanced Purchasing Agreements.<sup>16</sup> |
| EUR to USD exchange rate | World Bank<sup>44</sup> | Used for 2020. |
| Vaccine delivery cost of US\$ 3.7 per-person vaccinated | Global Dashboard for Vaccine Equity<sup>45</sup> | Due to the lack of consistent data and proxy cost data for COVID-19 vaccine delivery costs, a single value for all vaccines delivered was used. This value was applied to the vaccine coverage rates of each country, which were the vaccine coverage rates used in the modelling of COVID-19 vaccine impact, to obtain the total delivery costs.<sup>1</sup> |
| **Other** |  |  |
| World Bank income group classifications | World Bank<sup>46</sup> | Used 2022 classifications, which are based on 2021 data. |
| Vaccine coverage | Published literature | Proportion of the population with a full dose in the modelled countries by December 8, 2021. |

### Study perspective and horizon

The study follows a global payer perspective. Only true economic costs and benefits were considered, which excludes fiscal transfers between entities such as government vaccine purchases from manufacturers or intermediaries, including COVAX.^11^ Under this perspective, all costs and benefits associated with the vaccines are included, regardless of who incurred them.^12^ The time horizon includes COVID-19 vaccine development and delivery costs between 2020-2021, and the health and economic benefits generated by the vaccines delivered between 8 December 2020–8 December 2021 (including future life years lost from deaths in 2020–2021). All costs are reported in US$2021.

### Benefits

The health benefits generated from COVID-19 vaccines were evaluated in terms of life years (LYs) and quality-adjusted life years (QALYs) gained. LYs gained were determined by multiplying age group-specific average life expectancies for each country by the number of deaths averted, with a 3% discount rate applied to reflect time preferences for current health benefits. Undiscounted and discounted QALYs gained were estimated by combining averted QALY losses from infections, hospitalizations, and deaths. Further details are in Appendix B.

For the welfarist analysis, LYs gained were monetised by applying the value of statistical life years (VSLYs). Following recommendations from Robinson et al.,^13^ VSLYs were derived using country-specific income adjustments. While Robinson et al. only used discounted LYs, this analysis also includes an undiscounted approach, in which an undiscounted life expectancy and undiscounted LYs gained were used to calculate an undiscounted VSLY. For the extra-welfarist analysis, undiscounted and discounted QALYs gained were monetised by applying societal WTP thresholds, which are based on income-group dependent percentages of GDP per capita. Further details are in Appendix B.

Economic benefits generated during the first year of the COVID-19 vaccination programme were assessed from a societal perspective, by including both averted productivity losses and healthcare costs. Productivity costs averted due to reduced COVID-19 related absenteeism and deaths were calculated using the friction cost (FC) approach. This approach was chosen over the human capital approach to be conservative in estimating vaccine benefits; the FC approach assumes that sick or deceased workers are replaceable and only considers productivity costs incurred during the period when the position is unfilled (friction period). Healthcare costs averted were estimated by multiplying the number of hospitalisations averted by costs per hospitalisation. GDP deflator indices were used to control for inflation for costs that were reported in years other than 2021. Further details are in Appendix B. Since costs and benefits were only considered for one year, discounting was not applied to the costs. However, since life years gained extend beyond this first year, discounting was applied to adjust for preference for immediate benefits.

### Vaccine development and delivery costs

Development costs included costs from the pandemic’s start (2020), but excluded any costs related to the basic research that contributed to the vaccine development prior to the pandemic, as these were regarded as sunk costs. Vaccine development costs included both public and corporate investments into the research, development, and manufacturing of COVID-19 vaccines.^14–16^ Cost breakdowns are in Supplementary Table 1. Costs incurred in 2020 were converted to 2021 USD using GDP deflator indices.

Vaccine delivery costs reflect the costs incurred to deliver vaccines to individuals, including human resources and vaccine transport. A single vaccine delivery cost reflecting the average delivery cost for a fully vaccinated individual was used and multiplied by the number of fully vaccinated individuals.^17^

### Return-on-investment estimate

To determine the ROI, the monetised benefits were compared to the costs using the following general equation: ROI=(benefits–costs)/costs.^18^

Four ROI estimates were obtained, covering combinations of discounted/undiscounted and welfarist/extra-welfarist approaches.

### Sensitivity analyses

A probabilistic sensitivity analysis (PSA) was used to propagate parameter uncertainty to the ROI estimates. The PSA used Latin Hypercube Sampling (LHS) to generate 100 sets of values by sampling from the uncertainty distributions of the following parameters: (a) USA VSL value, (b) percentages of GDP used to calculate WTP thresholds for each World Bank income group, (c) QALY losses per infection, hospitalisation, and death, and (d) friction periods in HICs. We then used the 100 epidemic model fits previously generated in Watson et al. Each model fit represented a distinct set of epidemic parameters drawn from their respective uncertainty distributions. These outputs were randomly paired with the 100 sets of health-economic parameters sampled using LHS to capture uncertainty in both epidemiological and economic parameters. Median and 95% credible intervals for the combined 100 output sets were then calculated to determine the PSA results. Further details can be found in Appendix B. Partial rank correlation coefficients were calculated to assess the relationship between parameters and epidemiological model samples with reported uncertainties and their outcomes, and consequently the effects of parameter uncertainty on ROI estimates. Outcomes assessed in the PSA were monetized QALYs, friction costs and VSLYs.

### Statistical analysis

Analyses were conducted using R (version 2024.04.2+764). This paper was developed in alignment with the Consolidated Health Economic Evaluation Reporting Standards (CHEERS) 2022 guidelines (Appendix A).^19^

## Results

The total cost of COVID-19 vaccine development and delivery is estimated at $79.4 billion. Publicly announced public and non-profit investments into COVID-19 vaccine development between 2020-2021 are estimated at $13.2 billion. Advanced Purchase Agreements (APA) for COVID-19 vaccines totalled an estimated $42.7 billion. Corporate and private sector investments in development and manufacturing efforts are estimated at $14.1 billion. Additional manufacturing costs, excluding those included in public and corporate investments and APAs, totalled an estimated $389 million. With an estimated delivery cost of $3.70 per-person vaccinated, total delivery costs for all doses delivered in the first year of vaccination are estimated to be $8.94 billion.

From 8 December 2020 to 8 December 2021, COVID-19 vaccines are estimated to have generated 353 million (95%CrI 341–362) undiscounted LYs or 252 million (95%CrI 243–258) discounted LYs globally. LYs gained per-person vaccinated were greatest in high income countries (HICs) and lowest in low-income countries (LICs) (Supplementary Table 2). Vaccination was also estimated to have averted 1.50 billion (95%CrI 1.45–1.54) COVID-19 infections and 51.5 million (95%CrI 243–258) hospitalisations globally (Supplementary Table 3). We estimated that HICs and LICs experienced the greatest number of infections and hospitalisations averted per-person vaccinated, with relatively more infections averted per-person vaccinated in LICs.

Deaths, infections, and hospitalisations averted by COVID-19 vaccination generated an estimated 364 million (95%CrI 353–374) undiscounted QALYs globally. Applying a discount rate of 3% reduced this value to 263 million (95%CrI 254–270) QALYs gained. QALYs gained per-person vaccinated decreased with income group level (Supplementary Table 2).

Using a welfarist approach, it was estimated that COVID-19 vaccination generated economic value of $27.2 trillion (95%CrI 26.4–28.6). Applying a discount rate of 3% increased this value to $37.8 trillion (95%CrI 36.6–39.7). For both undiscounted and discounted VSLYs gained, HICs experienced the highest gains, while LICs experienced the lowest gains (Table 2). Consequently, the global VSLYs averted per-person vaccinated as a percentage of global GDPpc increased relative to each income group, driven primarily by high VSLYs from HIC combined with the lower GDPpc and larger populations in other income groups.

**Table 2.**
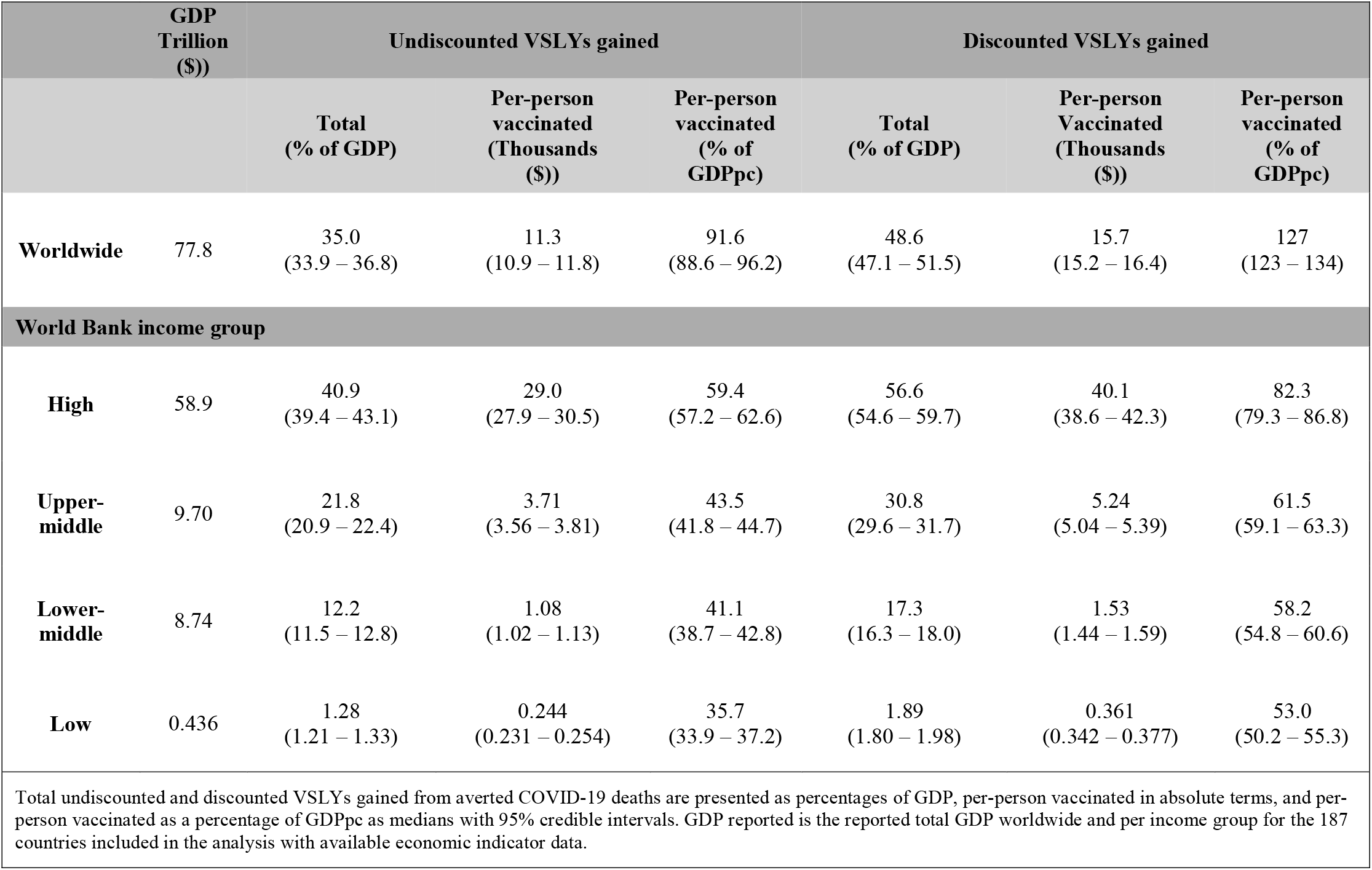
Estimates of health and economic benefits of COVID-19 vaccination using a welfarist framework through value of statistical life years gained expressed in three ways.

Using an extra-welfarist approach, COVID-19 vaccines generated $5.66 trillion (95%CrI 5.47–5.93) in economic value. After applying a discount rate of 3%, the QALYs gained decreased to $4.04 trillion (95%CrI 3.91–4.24). HICs generated the greatest savings from monetised QALYs in terms of dollars saved per-person vaccinated as a percentage of GDP, with LICs experiencing the lowest returns (Table 3).

Productivity costs saved from averted COVID-19 morbidity and mortality are estimated to be $564 billion (95%CrI 541–586), with LICs experiencing the greatest returns in dollars expressed as a percentage of GDPpc. COVID-19 vaccines were estimated to yield additional savings of $222 billion (95%CrI 213–229) in COVID-19-related healthcare costs. These savings in terms of a percentage of GDPpc were greatest in LICs and lowest in LMICs (Table 3).

**Table 3.**
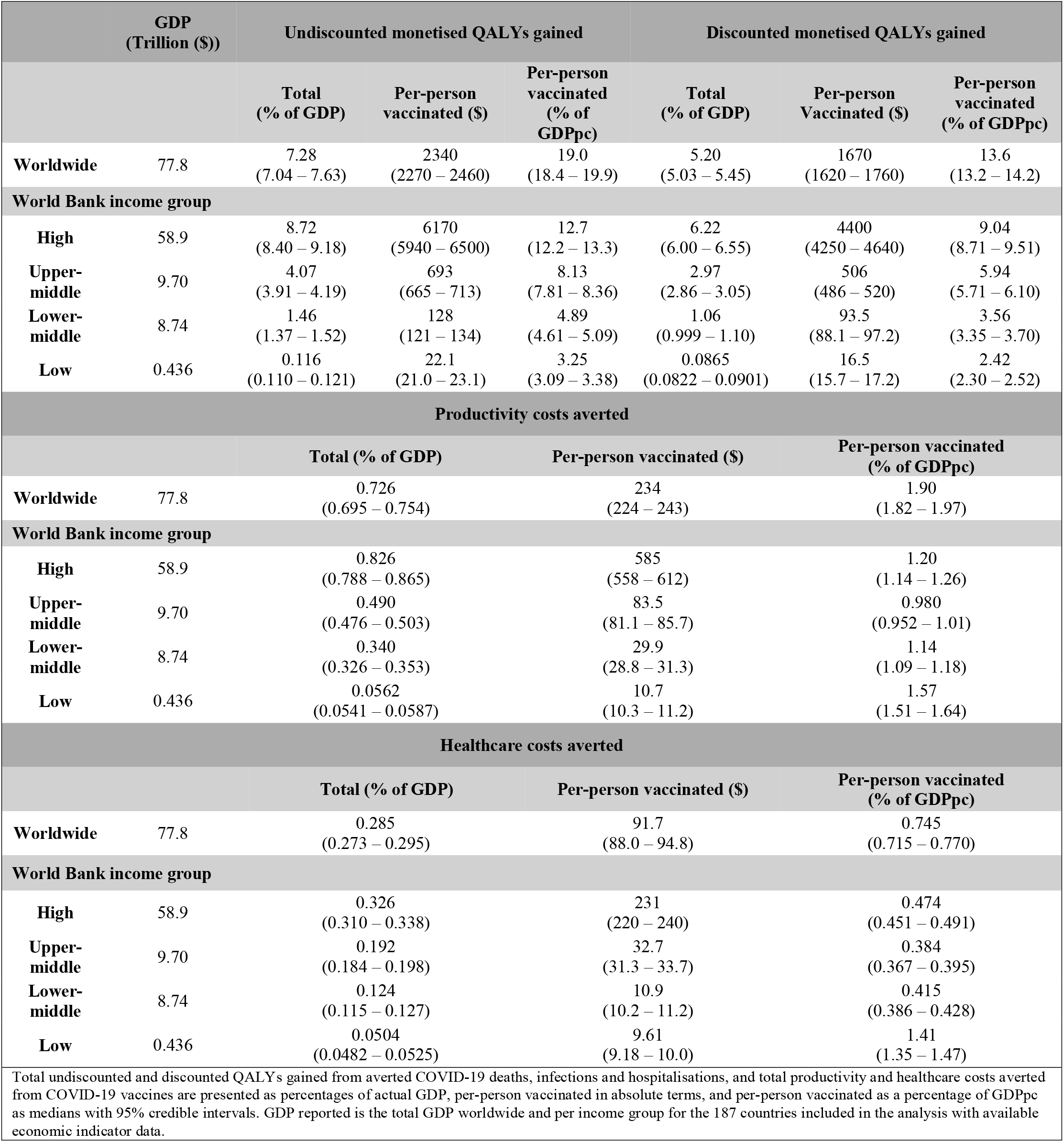
Estimates of monetised health and economic benefits of COVID-19 vaccines using an extra-welfarist approach, including monetized QALYs, and averted healthcare and productivity cost, expressed in three ways.

From an extra-welfarist approach, COVID-19 vaccination generated economic and health benefits valued at an estimated $6.44 trillion (95%CrI 6.22–6.75). Comparatively, under a welfarist approach, the monetized benefits are estimated at $27.2 trillion (95%CrI 26.4–28.6). When discounting was applied, the estimates under the two approaches are estimated to be $4.83 trillion (95%CrI 4.67–5.05) and $37.8 trillion (95%CrI 36.6–39.7), respectively. Global distributions of discounted monetized health and economic benefits can be seen in Figure 1 (undiscounted benefits in Supplementary Figure 1).

**Figure 1.**
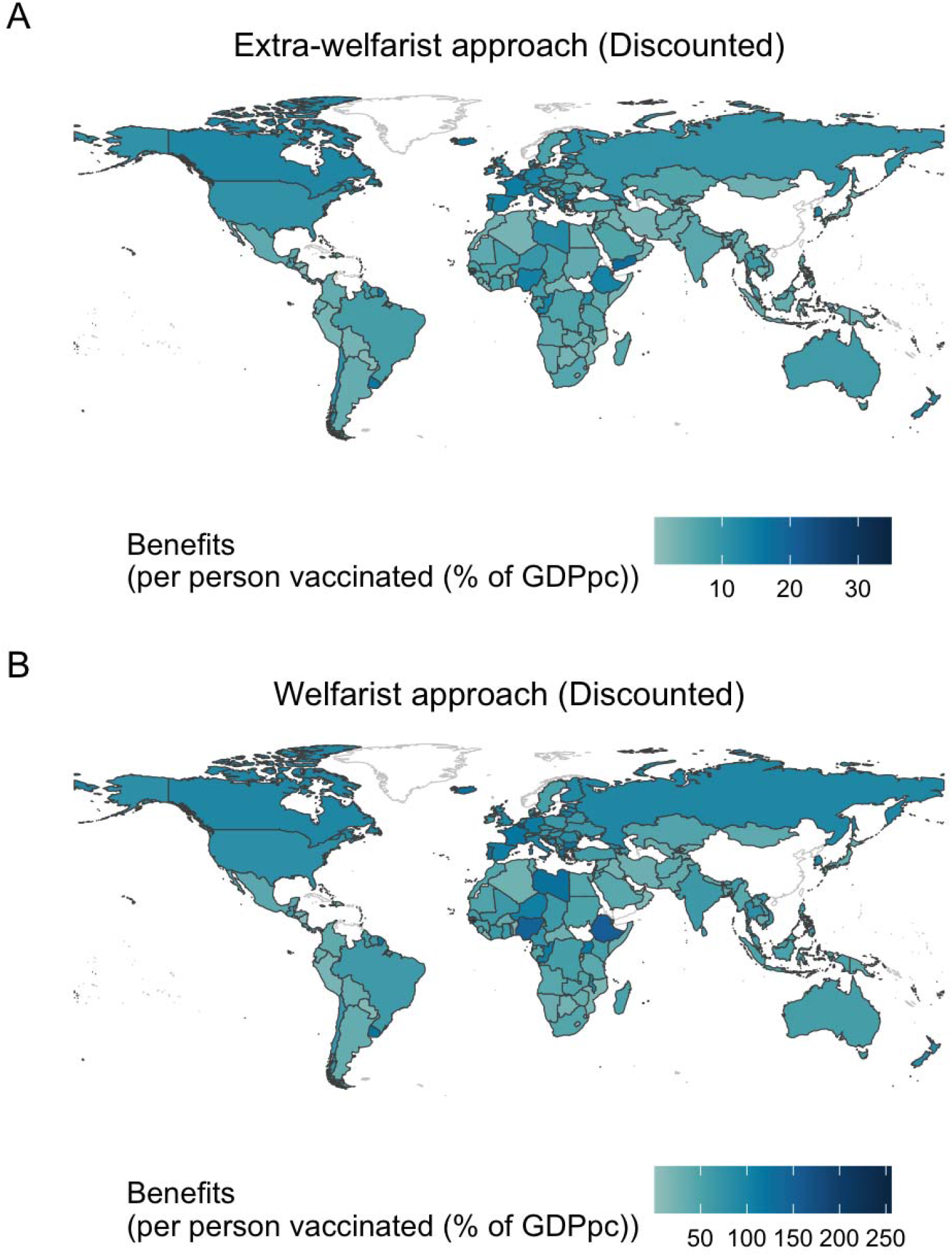
Global distribution of discounted monetized benefits from COVID-19 vaccination following extra-welfarist and welfarist approaches Figure 1A displays the global distribution of the sum of monetized benefits per-person vaccinated under the extra-welfarist approach, including monetized QALYs gained, and productivity and healthcare costs averted, after a discount rate of 3% was applied. These results are expressed as a percentage of GDP per capita across different countries. Figure 1B displays the global distribution of the sum of the monetized benefits per-person vaccinated under the welfarist approach, including VSLYs, expressed as a percentage of GDP per capita after a discount rate of 3% was applied. Countries with no data available are shown outlined in light grey with no colour fill.

Our results indicate an ROI well in excess of 1 for COVID-19 vaccines globally for the first year of the vaccination programme, under both approaches (Table 4). Based on the PSA, the welfarist approach produced markedly higher discounted ROI estimates (442 [95%CrI: 177–924]) compared to the extra-welfarist approach (62.2 [95%CrI: 25.0–83.1]). Across all approaches, the 95% CIs of the ROI exceeded 1 for the first year of the COVID-19 vaccination programme globally. Effects of parameter uncertainty on outcomes are found in Supplementary Figures 2-3. We found that WTP thresholds were the most influential parameter driving QALYs, according to the PRCC, showing a strong positive correlation with monetized QALY outcomes. For VSLYs, we found a positive correlation between VSL in the USA, deaths and hospitalisations averted with increased VSLYs, but a negative correlation for infections averted, likely reflecting residual variation in infections occurring among younger populations with limited mortality reflecting that after controlling for mortality, infections predominantly occurred in younger populations who contribute less to overall mortality.

**Table 4.** Return-on-investment estimates for the first year of the COVID-19 vaccination programme globally following the base case and probabilistic sensitivity analyses.

| | Undiscounted ROI (\$) | Discounted ROI (\$) |
| --- | --- | --- |
| **Base case analysis: Median (95% CI)** |  |  |
| Extra-welfarist approach | 80.2 (77.4 – 84.0) | 59.8 (57.8 – 62.7) |
| Welfarist approach | 342 (331 – 359) | 475 (460 – 499) |
| **Probabilistic sensitivity analysis: Median (95% CI)** |  |  |
| Extra-welfarist approach | 83.4 (31.6 – 113) | 62.2 (25.0 – 83.1) |
| Welfarist approach | 318 (127 – 665) | 442 (177 – 924) |

## Discussion

This study represents the first global ROI analysis for COVID-19 vaccines. In the first year of the vaccination programme, these vaccines were found to have a ROI far greater than 1 from both extra-welfarist and welfarist perspectives. Globally, for every dollar invested in vaccine development and delivery, the estimated returns ranged from $42.0 to $775. To our knowledge, this study provides the first economic evidence that the unprecedented, rapid investments made into COVID-19 vaccines paid off both in lives saved and long-term economic benefits. The overall findings were robust to changes in the approaches used (welfarist or extra-welfarist, discounted or undiscounted) and in various sensitivity analyses.

Our findings are consistent with other, more limited economic evaluations of COVID-19 vaccination, which also found that COVID-19 vaccines had high socioeconomic value on global, country and regional levels.^3,4,20–24^ The only other global analysis of COVID-19 vaccines’ value was conducted by Sevilla et al.^3^ This analysis had a much lower estimate of the number of lives saved by vaccination (4.1 million vs. 14-20 million from the model we used).^1^ The difference between the two studies largely results from their reliance on reported COVID-19 case data (which is known to underestimate true burden in countries with weaker vital registration systems).^25^ Differences are also due to the regression approach used in Sevilla et al., compared to the compartmental transmission model used by Watson et al.^1^ Despite the health impact difference, Sevilla et al. estimated that COVID-19 vaccines generated $5.2 trillion in value in health gains and GDP impact, similar to our equivalent extra-welfarist value.^3^ The similarity between the two value figures is driven by the inclusion of vaccine impact on GDP in Sevilla et al., which was estimated through regression independent of the health impact estimates.

Our ROI of COVID-19 vaccination exceeds the already high ROIs of routine vaccination programmes in LMICs in Ozawa et al.^26^ That study reported an ROI of 22.2 per dollar invested in a range of vaccines (measles, rubella, rotavirus, pneumococcal and HPV vaccines) under a cost-of-illness approach (similar to our extra-welfarist approach, but without monetising QALYs), and 51.8 per dollar invested under a VSL approach (similar to our welfarist approach). Differences in estimates can partly be attributable to methodological differences (lack of QALY monetisation in the cost-of-illness approach used by Ozawa) and setting (Ozawa restricted to LMICs, where productivity costs are lower). However, our higher ROI values may also reflect the uniquely valuable contribution that COVID-19 vaccines made in the first year of a severe pandemic.

Despite our ROI estimates being greater than 1, the study reveals significant disparities in vaccine benefits across income groups. HICs gained the greatest economic benefits due to early access, higher coverage, and faster rollouts, especially before the Delta variant’s emergence in mid-2021.^1^ The older populations of HICs also contributed to greater vaccine benefit, as the per-person risk of severe outcomes was higher. In contrast, many LMICs had not yet begun vaccinating most of their populations during the Delta wave (mid-2021), and some LMICs (i.e., many countries in sub-Saharan Africa) had not even been able to achieve high vaccine coverage during the Omicron wave (late 2021). Hence, the value of vaccination was diminished as their populations had already received immunity through natural infection instead.^1^ Earlier global access to COVID-19 vaccines would have led to substantial benefits, especially in LMICs, as reported by Barnsley et al.^24^ Additionally, HICs and upper-middle income countries (UMICs) could afford more efficacious vaccines (e.g., mRNA vaccines) which conferred greater health and economic benefits.^1^ In contrast, LICs saw smaller total economic gains; however, their health benefits per-person vaccinated were relatively high as the low vaccine coverage targeted the most vulnerable. LICs also experienced the highest averted healthcare and productivity costs per-person vaccinated relative to GDPpc, emphasizing the profound economic benefits vaccines had on these countries despite their late introduction, and the importance of investments into vaccination programs in LICs to enhance health equity and global economic stability.

Disparities between countries highlight the need for more equitable vaccine distribution strategies in global health crises, particularly in the early stages. While the COVID-19 Vaccines Global Access (COVAX) facility aimed to create a fair allocation mechanism for COVID-19 vaccines, several challenges, including constrained and delayed vaccine supply to COVAX, prevented COVAX from reaching its target of 20% coverage in each country.^1^ Had COVAX reached these goals, the benefits of vaccination would have been more equitably distributed as LMICs would have had access to vaccines sooner. The recently signed Pandemic Treaty promises LMICs access to vital technologies, including vaccines, in subsequent pandemics, although it lacks legally binding enforcement mechanisms and excludes participation by key countries such as the United States.^8^ The most secure route may be to make vaccines a global public good during pandemics by removing restrictions on vaccine intellectual property and spreading vaccine production capacity to more countries.^27,28^ Further actions to improve vaccine equity could include increased investment from HICs in vaccine development partnerships involving governments, universities, and companies and their counterparts in LMICs. These partnerships will increase capacity in LMICs to help ensure equity and maintenance of a worldwide collaboration network^29^, with potential long-term economic benefits across all partners.

While this study provides important insights, it has limitations. Our results rely on previous analyses of COVID-19 vaccination impact, which is estimated based on a counterfactual modelling approach.^1^ This approach is common practice in vaccine impact studies. However, it relies on simplifying assumptions around the counterfactual, such as assuming the level of public health and social restrictions, emergence of variants of concern, and human behaviour and mixing remain the same as observed during the COVID-19 pandemic regardless of vaccine coverage. Further, this study only evaluates the ROI for the first year of COVID-19 vaccines. Although this was the most important year for vaccine impact as well as the year with the greatest vaccine development costs, incorporating additional benefits and costs that have occurred since would likely make the ROI estimates even more favourable. This includes booster doses (and continued rollout of primary courses) in response to the Omicron variant and its sub-lineages, which are likely to have also averted a substantial number of hospitalisations and deaths, as well as the health and economic impacts of long COVID-19. Other limitations include the lack of precise data and uncertainty estimates around vaccine manufacturing and delivery costs.

This study provides strong evidence to support investments made into COVID-19 vaccine development and delivery. The findings emphasize the critical economic value of vaccines in safeguarding public health and preserving global economic stability, which is important to establish at a time of reduced vaccine funding, particularly in LMICs, and heightened scepticism globally about vaccine value. Our findings highlight the value of investment in vaccine research and development before and during pandemics, as well as the importance of equitable vaccine access and distribution. Ensuring vaccination program benefits are accessible to all populations, regardless of income level, is essential for enhancing global health equity and preparedness against future pandemics.

## Data Availability

All data produced in the present study are available upon reasonable request to the authors.

## Authors’ contributions

HB, MJ and OJW conceived the study with input from GB. HB conducted the analysis with the assistance of OJW and GB. HB, MJ and OJ produced the first draft of the manuscript and have accessed and verified the underlying data. All authors read, contributed to, and approved the final draft. All authors had full access to all the data in the study and had final responsibility for the decision to submit for publication.

## Acknowledgments

This study is funded by the National Institute for Health and Care Research (NIHR) Health Protection Research Unit in Modelling and Health Economics, a partnership between the UK Health Security Agency, Imperial College London and LSHTM (grant code NIHR200908). The views expressed are those of the author(s) and not necessarily those of the NIHR, UK Health Security Agency or the Department of Health and Social Care. OJW was supported by an Imperial College and Eric and Wendy Schmidt AI in Science Fellowship, a program of Schmidt Sciences, Centre funding from the UK Medical Research Council (reference MR/X020258/1), and funding from Community Jameel. GB was funded by the Medical Research Council (MR/W006677/1).

