## Supplementary Materials for "The global return-on-investment of COVID-19 vaccines in the first year of the vaccination programme"

**Appendix A: CHEERS Checklist**

| **Topic** | **No.** | **Item** | **Location where item is reported** |
| --- | --- | --- | --- |
| **Title** |  |  |  |
|  | 1 | Identify the study as an economic evaluation and specify the interventions being compared. | Page 1 |
| **Abstract** |  |  |  |
|  | 2 | Provide a structured summary that highlights context, key methods, results, and alternative analyses. | Page 2 |
| **Introduction** |  |  |  |
| **Background and objectives** | 3 | Give the context for the study, the study question, and its practical relevance for decision making in policy or practice. | Page 3 |
| **Methods** |  |  |  |
| **Health economic analysis plan** | 4 | Indicate whether a health economic analysis plan was developed and where available. | Page 3 |
| **Study population** | 5 | Describe characteristics of the study population (such as age range, demographics, socioeconomic, or clinical characteristics). | Page 4 |
| **Setting and location** | 6 | Provide relevant contextual information that may influence findings. | Page 4 |
| **Comparators** | 7 | Describe the interventions or strategies being compared and why chosen. | Page 4 |
| **Perspective** | 8 | State the perspective(s) adopted by the study and why chosen. | Page 4 |
| **Time horizon** | 9 | State the time horizon for the study and why appropriate. | Page 4 |
| **Discount rate** | 10 | Report the discount rate(s) and reason chosen. | Page 4 and Appendix B |
| **Selection of outcomes** | 11 | Describe what outcomes were used as the measure(s) of benefit(s) and harm(s). | Page 6 |
| **Measurement of outcomes** | 12 | Describe how outcomes used to capture benefit(s) and harm(s) were measured. | Page 6 and Appendix B |
| **Valuation of outcomes** | 13 | Describe the population and methods used to measure and value outcomes. | Page 6 and Appendix B |
| **Measurement and valuation of resources and costs** | 14 | Describe how costs were valued. | Page 6 |
| **Currency, price date, and conversion** | 15 | Report the dates of the estimated resource quantities and unit costs, plus the currency and year of conversion. | Page 4 |
| **Rationale and description of model** | 16 | If modelling is used, describe in detail and why used. Report if the model is publicly available and where it can be accessed. | Page 4 and Appendix B |
| **Analytics and assumptions** | 17 | Describe any methods for analysing or statistically transforming data, any extrapolation methods, and approaches for validating any model used. | Page 7 |
| **Characterising heterogeneity** | 18 | Describe any methods used for estimating how the results of the study vary for subgroups. | Appendix B |
| **Characterising distributional effects** | 19 | Describe how impacts are distributed across different individuals or adjustments made to reflect priority populations. | Appendix B |
| **Characterising uncertainty** | 20 | Describe methods to characterise any sources of uncertainty in the analysis. | Page 7 |
| **Approach to engagement with patients and others affected by the study** | 21 | Describe any approaches to engage patients or service recipients, the general public, communities, or stakeholders (such as clinicians or payers) in the design of the study. | N/A |
| **Results** |  |  |  |
| **Study parameters** | 22 | Report all analytic inputs (such as values, ranges, references) including uncertainty or distributional assumptions. | Page 4-6 |
| **Summary of main results** | 23 | Report the mean values for the main categories of costs and outcomes of interest and summarise them in the most appropriate overall measure. | Page 7-13 |
| **Effect of uncertainty** | 24 | Describe how uncertainty about analytic judgments, inputs, or projections affect findings. Report the effect of choice of discount rate and time horizon, if applicable. | Page 13-14 |
| **Effect of engagement with patients and others affected by the study** | 25 | Report on any difference patient/service recipient, general public, community, or stakeholder involvement made to the approach or findings of the study | N/A |
| **Discussion** |  |  |  |
| **Study findings, limitations, generalisability, and current knowledge** | 26 | Report key findings, limitations, ethical or equity considerations not captured, and how these could affect patients, policy, or practice. | Page 15-16 |
| **Other relevant information** |  |  |  |
| **Source of funding** | 27 | Describe how the study was funded and any role of the funder in the identification, design, conduct, and reporting of the analysis | Page 2 and 17 |
| **Conflicts of interest** | 28 | Report authors conflicts of interest according to journal or International Committee of Medical Journal Editors requirements. | N/A |

*From:* Husereau D, Drummond M, Augustovski F, et al. Consolidated Health Economic Evaluation Reporting Standards 2022 (CHEERS 2022) Explanation and Elaboration: A Report of the ISPOR CHEERS II Good Practices Task Force. Value Health 2022;25. <doi:10.1016/j.jval.2021.10.008>

**Appendix B: Methods**

**Data collection**

To accurately assess the impact of COVID-19 vaccinations, our study relied on data sourced from a comprehensive global analysis of the impact of COVID-19 vaccination by Watson et al. This analysis provided estimates of the number of infections, hospitalisations, and deaths averted due to COVID-19 vaccination efforts worldwide for seventeen discrete age groups (0-5, 5-10, ..., 75-80, 80+) (Table 1). These public health outcomes (averted infections, hospitalisations and deaths) reported in Watson et al. were estimated by fitting a mechanistic transmission model of COVID-19 transmission and vaccination using novel fitting process-incorporating uncertainty in various epidemiological parameters. This model was fit to the time series of excess deaths recorded during the pandemic in each country. The authors estimated the public health outcomes that were averted due to vaccination by comparing the fitted model against a counterfactual scenario in which COVID-19 vaccinations were not administered, but all other public health interventions, such as NPIs, remained in place.

**Estimating health benefits**

For this health economic study, the public health outcomes from Watson et al. (2022) were used(1). To capture the uncertainty in these outcomes, we drew 100 samples from the posterior distribution for each country analysed. These public health outcomes include COVID-19 deaths, infections, and hospitalisations averted by the vaccines, aggregated by age group per country. The median and 95% credible intervals (CrI) for these outcomes within each country were estimated, with the 95% CrI calculated as the 95% central percentile from the 100 draws. Additionally, for aggregated estimates across different World Bank income groups, a sampling strategy where one draw was sampled from each country’s posterior distribution was employed. These individual draws from each country were subsequently summed to represent the total averted outcomes for that specific income group in a single simulation. This process was repeated 100 times to ensure that draws between countries were uncorrelated, with the resulting 100 draws used to calculate the median and 95% credible intervals for each health outcome.

When using the results from the Bayesian infectious disease model to calculate health economic outputs, the medians and 95% credible intervals were multiplied by fixed parameters. The fixed parameters include those in Table 1 of the main text.

**Averted infections, hospitalisation and life years gained**

The public health outcomes from Watson et al. (1) were used to report the infections and hospitalisations averted by COVID-19 vaccines.

Using the number of deaths averted by the vaccines and life expectancies, the number of LYs gained was estimated. The LYs gained were based on 5-year-band age-groups, beginning with 0-5 and ending with 80+. The undiscounted LYs gained for each age-group was calculated by multiplying each group’s average life expectancy by the number of deaths in that group. A discount rate of 3% was then applied to reflect the time preferences for health benefits to determine the discounted LYs gained(2–4). No female, male or health status differences were considered.

**Quality-adjusted life years gained**

A QALY is a health outcome measure that simultaneously captures changes in health-related quality of life, and changes in the duration of life due to premature deaths(5). This is calculated based on health state utilities on a scale in which a value of 1.0 is full health and a value of zero is death(6). The utilities assigned to health states are preference weights, based on time trade-offs where respondents compare between existing in a certain health state for a longer period of time before death, and a more preferred health state but for a shorter period of time (7). These health utilities are then multiplied by changes in life duration to determine the number of QALYs in a particular scenario and compared between scenarios to estfimate the QALYs gained or lost by a particular intervention. The number of QALYs gained from COVID-19 vaccines was estimated from the averted QALY losses associated with COVID-19 infections, hospitalisations, and deaths (i.e. the difference in these outcomes between vaccination and non-vaccination scenarios). The number of QALY lost from a single COVID-19 case, hospitalisation or death was obtained from an study by Basu and Gandhay (2021), where a cohort-based probabilistic stimulation model using epidemiological data provided by the United States’ Centre for Disease Control and Prevention was used to quantify the QALY losses associated with COVID-19 disease states(8). The quality-of-life weights for the disease states were obtained from patients with severe influenzas and H1N1 using the EQ-5D(8). Equations 1 and 2 represent the calculations used to calculate the estimated QALYs gained.

**Equation 1: Estimating QALYs gained by COVID-19 vaccination from averted infections**

Infection QALYs gained = – (QALY loss for COVID-19 infections) * averted infections

**Equation 2: Estimating QALYs gained by COVID-19 vaccination from averted hospitalisations**

Hospitalisation QALYs gained = – (QALY loss for COVID-19 hospitalisations) * averted hospitalisations

The number of QALYs associated with a COVID-19 death is a combination of the LYs lost due to premature death, and the non-fatal QALYs lost due to the acute COVID-19 episode leading to death. The way undiscounted and discounted QALYs gained by COVID-19 vaccination from averted deaths is calculated is shown in Equation 3 below.

**Equation 3: Estimating undiscounted QALYs gained by COVID-19 vaccination from averted deaths**

(Un)discounted non-fatal QALYs gained = -(Un)discounted life years lost - (non-fatal QALYs lost from a COVID-19-associated death x number of averted COVID-19 deaths)

The total (un)discounted QALYs gained from vaccination was then estimated as the sum of the QALYs gained from averted hospitalisations, infections and deaths, with the latter quantity based on (un)discounted LYs.

**Monetizing health benefits**

**Value of statistical life years**

The ‘value of a statistical life’ (VSL) quantifies the monetary value individuals place on small changes in mortality risk. VSLs are derived from stated and revealed preferences of individuals’ willingness to pay (WTP) for marginal reduction in their current mortality risk and are frequently used to evaluate the benefits of interventions that reduce mortality risk(9). Stated preferences are gathered from responses to surveys with carefully structured, hypothetical questions about an individual’s WTP for a lowered risk of death or disability. In contrast, revealed preferences are based on individuals’ observed behaviour gathered from labour market data. These revealed preferences represent the trade-offs between increased risk of morbidity or mortality for higher wages(9). In our analysis, we used reported VSLs to determine value of statistical life years (VSLYs) gained from averting COVID-19 deaths via vaccination. VSLYs represent an individual’s WTP for one-year gains in life expectancy(10). Compared to VSLs, VSLYs better reflect the long-term benefits of vaccination on health as VSLYs capture the impact of vaccination on providing additional life years(10). To produce global estimate of VSLYs, a similar methodology was adapted from Robinson et al. (2019)(11). VSLs for each country were estimated using a USA reference value. The reference VSL was multiplied by the ratio of gross national income per capita (GNIPC) between the country of interest and the USA to the power of the income elasticity (Equation 5)(11). GNIPC refers to the sum of the economic activities undertaken by citizens and firms of a country divided by the population(2). Income elasticity is the sensitivity of VSL changes in response to changes in income. An income elasticity of 1 was used, as recommended by Viscusi and Masterman (2018), given that it is a tractable value and their income elasticity analyses failed to reject the hypothesis that international income elasticity was equal to 1(12), i.e. a 10% increase in GNIPC results in a 10% increase in VSL.

**Equation 4: Country-specific VSL estimates from USA reference value**

$${VSL}_{x}=\left( \frac{{VSL}_{USA}*{GNIPC}_{x}}{{GNIPC}_{USA}} \right)^{E}$$

The derived VSLs were used to calculate VSLYs, based on age group-dependent life expectancies and the distribution of the population amongst age groups. Following Robinson et al. (2021), the VSL estimate was divided by a population-weighted discounted life expectancy, where $l_{g}$ is the discounted LYs remaining per age group and $N_{g}$ is the number of people per age group, to get the value of a single year of statistical life (VLY)(11). To get the total VSLYs gained, the VLY was multiplied by the number of discounted LYs gained (Equation 6). While Robinson et al. (2021) only used discounted LYs in their VSLY calculations, this analysis also includes an undiscounted approach, in which undiscounted life expectancy and undiscounted LYs gained were used to calculate undiscounted VSLYs following the same approach. Using an undiscounted approach assumes that individuals do not discount future LYs in their states and revealed preferences when determining their WTP, and therefore value current and future LYs equally.

**Equation 5: VSLY estimates using population-weighted life expectancy, VSL estimates and deaths averted per age group**

$$VLY=\frac{VSL}{\left( \frac{\sum_{g} N_{g}l_{g}}{\sum_{g} Ng} \right)}$$

$$VSLY=VLY*l_{g}averted$$

**Monetized QALYs gained**

To monetize the QALYs gained from COVID-19 vaccines, the QALYs gained were multiplied by societal WTP thresholds, which represent what society would pay for improvements in individuals’ health(2). Most countries do not have explicit WTP thresholds, so they were estimated by multiplying the country’s GDP per capita (GDPpc) by a specified percentage, depending on which income group the country belongs to. These percentages were obtained from Pichon-Riviere et al. (2023), who estimated cost-effectiveness thresholds for 174 countries, based on the economic capacities and opportunity costs of healthcare spending in each country income group. For example, to determine Canada’s WTP threshold, which is a high-income country (HIC), its GDPpc would be multiplied by 0.68, and to determine Uganda’s WTP thresholds, a low-income country (LIC), its GDPpc would be multiplied by 0.24. The WTP thresholds were then multiplied by the total QALYs gained from averted infections, hospitalisations, and deaths. This process was completed to obtain two estimates: undiscounted total monetized QALYs gained, and discounted total monetized QALYs gained (Equation 7 and 8).

**Equation 6: Estimating total monetized undiscounted QALYs gained**

Total monetized (un)discounted QALYs gained = (Equation 1 + Equation 2 + Equation 3) * % GDPpc

**Estimating economic benefits**

**Productivity costs**

COVID-19 morbidity and mortality caused significant productivity losses affecting the economies of countries globally. Productivity loss averted due to morbidity and mortality reductions is an important indirect benefit of COVID-19 vaccines(14). The costs of productivity losses averted by COVID-19 vaccines from COVID-19-related absenteeism for non-fatal cases and COVID-19 deaths were estimated using the friction cost (FC) approach. Through this approach, the indirect benefits of COVID-19 vaccines on productivity can be estimated. The FC method quantifies the costs incurred to the firm when workers are sick and cannot work or die and must be replaced. For unpaid workers, the opportunity cost approach was undertaken, in which the time spent on unpaid work is based on the value of spending this time in an alternative capacity (i.e. as a paid worker)(15).

**Non-fatal cases**

For non-fatal cases, the productivity costs incurred refer to the productivity loss during the time in which the worker was sick and unable to work. The productivity loss was valued by an individual’s average income per day multiplied by the duration of absenteeism. GDPpc was used as a proxy for average income and then divided by 365.25 to obtain the average income per day(16). This value was then multiped by the average absenteeism duration to determine COVID-19-related illness productivity losses. The absenteeism duration was based on the duration of COVID-19-related infections and hospitalisations. To obtain the productivity loss averted, the above value was multiplied by the number of cases averted (Equation 8). This process was completed for both hospitalisations and infections averted, using their respective durations, and then summed together to get the overall productivity costs averted for non-fatal cases.

**Equation 8: Productivity costs averted from averted COVID-19 morbidity by COVID-19 vaccines**

Productivity costs averted = (GDPpc / 365.25) * days absent * cases averted

**Fatal cases**

When someone dies from COVID-19, it can be assumed that there is a friction period where someone is not in that role, and therefore a cost is incurred to the firm(14). These productivity costs were determined by multiplying the friction period duration (in days) by the average daily income. GDPpc divided by 365.25 was used as a proxy for average daily income(16). To determine the costs averted for fatal cases, this above value was multiplied by the number of deaths averted (Equation 9).

**Equation 9: Productivity costs averted from averted COVID-19 mortality by COVID-19 vaccines**

Productivity costs averted = (GDPpc / 365.25) * friction period * deaths averted

The sum of the friction costs gained from non-fatal and fatal cases was calculated and used in the analysis.

**Healthcare costs**

Another economic benefit of COVID-19 vaccines was the attributable reductions in healthcare costs due to averted hospitalisations. For low- and middle-income countries (LMICs), healthcare costs per day for COVID-19-related hospitalisations were obtained from Torres-Rueda et al. (2021), which reported the data in US$2019. To inflate this to US$2021, the data was multiplied by a ratio of US$2021 GDP deflator index to the US$2019 GDP deflator index (17). To obtain the total healthcare cost per hospitalisation, the cost per day was multiplied by the hospitalisation duration. This cost was then multiplied by the number of hospitalisations averted to get the healthcare costs saved. For HICs, unit costs for hospitalisation were obtained from WHO-CHOICE and reported in 2010 International Dollars (I$). The figures used were “Cost per inpatient bed day by hospital level (without drugs)” values for primary hospitals. To convert it to US$2021, given that I$1 reflects the comparable amount that US$1 could buy in the USA in the country and currency of interest, the data was multiplied by a ratio of 2021 to 2010 GDP deflator indexes to account for inflation(17). Cost per hospitalisation was then obtained by multiplying the cost by the hospitalisation duration. To get the total costs averted, the cost per hospitalisation was multiplied by the total number of hospitalisations averted (Equation 10).

**Equation 10: Estimating healthcare costs averted from averted COVID-19 hospitalisations by COVID-19 vaccines**

Healthcare costs = costs per hospitalisation * duration * hospitalisations averted

**Probabilistic sensitivity analyses**

A probabilistic sensitivity analysis (PSA) was used to propagate parameter uncertainty to the ROI estimates. The PSA used Latin Hypercube Sampling (LHS) to generate 100 sets of values for parameters with reported uncertainties. The USA VSL value used in the VSLY calculations was modeled using a gamma distribution, with shape and scale parameters derived from the reported mean and standard deviation. Percentages of GDP used to obtain WTP thresholds were modeled using beta distributions, with shape parameters estimated by optimization between theoretical and empirical median and interquartile range values from the literature. QALY losses were also modeled using beta distributions, with shape parameters derived from reported mean values and 95% confidence intervals. Friction periods of HICs were modeled using a normal distribution with the reported mean and standard deviation from the literature. The PSA samples were randomly matched with the COVID-19 vaccine modelling samples and other economic outcomes, including healthcare costs, by calculating the median and 95% credible intervals of the draws to determine the uncertainty in the ROI estimates.

**Supplementary Table 1**. Vaccine research and development costs, including public funding, non-profit funding, corporate investments and manufacturing costs

|  | Value (millions) | Currency and year | Source |
| --- | --- | --- | --- |
| Public and non-profit funding for research, development and production (includes some manufacturing costs) | | | |
|  | 2100 | USD, 2020 | (18) |
|  | 2100 |  |  |
|  | 1700 |  |  |
|  | 1500 |  |  |
|  | 957 |  |  |
|  | 445 |  |  |
|  | 530 |  |  |
|  | 348 |  |  |
|  | 142 |  |  |
|  | 137 |  |  |
|  | 107 |  |  |
|  | 15 |  |  |
|  | 14 |  |  |
|  | 9 |  |  |
|  | 4 |  |  |
|  | 3 |  |  |
|  | 0.70 | USD, 2020 | (19) |
|  | 86.89 |  |  |
|  | 15.01 |  |  |
|  | 17.49 |  |  |
|  | 0.23 |  |  |
|  | 42.03 |  |  |
|  | 5.00 |  |  |
|  | 9.80 |  |  |
|  | 2.98 |  |  |
|  | 2.59 |  |  |
|  | 0.70 |  |  |
|  | 0.75 |  |  |
|  | 0.43 |  |  |
|  | 0.61 |  |  |
|  | 0.39 |  |  |
|  | 8.20 |  |  |
|  | 25.88 |  |  |
|  | 1.50 |  |  |
|  | 2.11 |  |  |
|  | 2.11 |  |  |
|  | 0.70 |  |  |
|  | 0.09 |  |  |
|  | 0.39 |  |  |
|  | 0.84 |  |  |
|  | 347.55 |  |  |
|  | 454.31 |  |  |
|  | 85.30 |  |  |
|  | 25.88 |  |  |
|  | 115.85 |  |  |
|  | 23.00 |  |  |
|  | 271.76 |  |  |
|  | 250.00 |  |  |
|  | 7.00 |  |  |
|  | 1.58 |  |  |
|  | 132.07 |  |  |
|  | 20.00 |  |  |
|  | 1.50 |  |  |
|  | 0.52 |  |  |
|  | 13.49 |  |  |
|  | 3.48 |  |  |
|  | 1.41 |  |  |
|  | 0.7 |  |  |
|  | 0.7 |  |  |
|  | 0.51 |  |  |
|  | 0.17620575 |  |  |
|  | 0.070483 |  |  |
|  | 1.41 |  |  |
|  | 237.00 |  |  |
|  | 162.19 |  |  |
|  | 3.88 |  |  |
|  | 3.02 |  |  |
|  | 27.00 |  |  |
|  | 4.90 |  |  |
|  | 8.30 |  |  |
|  | 5.70 |  |  |
|  | 2.36 |  |  |
|  | 4.00 |  |  |
|  | 5.79 |  |  |
|  | 0.30 |  |  |
|  | 0.41 |  |  |
|  | 10.00 |  |  |
|  | 10.81 |  |  |
|  | 7.05 |  |  |
|  | 57.93 |  |  |
|  | 2.20 |  |  |
|  | 0.94 |  |  |
|  | 1.41 |  |  |
|  | 0.75 |  |  |
|  | 0.75 |  |  |
|  | 0.54 |  |  |
|  | 0.56 |  |  |
|  | 0.45 |  |  |
|  | 0.75 |  |  |
|  | 2.47 |  |  |
|  | 150 |  |  |
|  | 6.90 |  |  |
|  | 29.12 |  |  |
|  | 17.26 |  |  |
|  | 11.48 |  |  |
|  | 8.25 |  |  |
|  | 9.01 |  |  |
|  | 11.26 |  |  |
|  | 4.63 |  |  |
|  | 1.16 |  |  |
|  | 2.15 |  |  |
|  | 8.00 |  |  |
|  | 1.30 |  |  |
|  | 115.85 |  |  |
|  | 7.05 |  |  |
|  | 5.29 |  |  |
|  | 86.89 |  |  |
|  | 0.49 |  |  |
|  | 0.28 |  |  |
|  | 1.06 |  |  |
|  | 0.14 |  |  |
|  | 0.34 |  |  |
|  | 2.32 |  |  |
|  | 1.00 |  |  |
|  | 11.90 |  |  |
|  | 23.94 |  |  |
|  | 3.16 |  |  |
|  | 38.03 |  |  |
|  | 9.91 |  |  |
|  | 1.41 |  |  |
|  | 1.74 |  |  |
|  | 144.96 |  |  |
|  | 21.95 |  |  |
|  | 2.90 |  |  |
|  | 2.85 |  |  |
|  | 1.95 |  |  |
|  | 0.02 |  |  |
|  | 0.10 |  |  |
|  | 0.12 |  |  |
|  | 21.55 | USD, 2021 | (19) |
|  | 139.02 |  |  |
|  | 4.8 |  |  |
|  | 14.2 |  |  |
|  | 33 |  |  |
|  | 32 |  |  |
|  | 132.4 |  |  |
|  | 1.06 |  |  |
| Advanced purchasing agreements | | | |
|  | 275 | USD, 2020 | (19) |
|  | 175 |  |  |
|  | 469.09 |  |  |
|  | 750 |  |  |
|  | 40 |  |  |
|  | 1060 |  |  |
|  | 250 |  |  |
|  | 120 |  |  |
|  | 4.3 |  |  |
|  | 12.47 |  |  |
|  | 942.65 |  |  |
|  | 1700 |  |  |
|  | 4500 |  |  |
|  | 2880 |  |  |
|  | 11340 |  |  |
|  | 352.5 |  |  |
|  | 84 |  |  |
|  | 5972.48 |  |  |
|  | 2790 |  |  |
|  | 858 |  |  |
|  | 2200 | USD, 2021 | (19) |
| Manufacturing costs | | | |
|  | 204 | USD, 2020 | (19) |
|  | 99 | USD, 2021 |  |
|  | 77 | USD, 2021 |  |
| Corporate and private sector investments | | | |
|  | 11000 | EUR, 2020 | (20) |
|  | 1.16 | USD, 2020 | (19) |
|  | 500 |  |  |
|  | 0.33 |  |  |
|  | 0.33 |  |  |
|  | 1.01 |  |  |
|  | 7.50 |  |  |
|  | 7.50 |  |  |

**Supplementary Table 2.** Estimates of undiscounted and discounted life years and QALYs gained from averted COVID-19 morbidity and mortality by COVID-19 vaccines

|  | **Vaccine coverage (%)** | **Total**  **(million)** | | **Per 1000 people vaccinated** | **Total**  **(million)** | | **Per 1000 people vaccinated** |
| --- | --- | --- | --- | --- | --- | --- | --- |
|  |  | **Undiscounted LYs gained** | | | **Discounted LYs gained** | | |
| **Worldwide** | 38.8 | 353  (341 – 362) | | 146  (141 – 150) | 252  (243 – 258) | | 104  (101 – 107) |
| **World Bank income group** | | | | | | | |
| **High** | 68.8 | 147  (142 – 155) | | 177  (171 – 187) | 104  (100 – 109) | | 125  (120 – 132) |
| **Upper-middle** | 50.1 | 75.1  (72.3 – 77.3) | | 132  (127 – 136) | 53.9  (51.9 – 55.4) | | 94.5  (91.0 – 97.2) |
| **Lower-middle** | 29.8 | 128  (120 – 133) | | 129  (121 – 134) | 91.7  (86.0 – 95.1) | | 92.6  (86.8 – 95.9) |
| **Low** | 3.57 | 2.70  (2.57 – 2.81) | | 118  (113 – 123) | 2.00  (1.89 – 2.06) | | 86.8  (82.6 – 90.2) |
|  | | **Undiscounted QALYs** | | | **Discounted QALYs** | | |
| **Worldwide** | 38.8 | 364  (353 – 374) | 151  (146 – 155) | | 263  (254 – 270) | 109  (105 –112) | |
| **World Bank income group** | | | | | | | |
| **High** | 68.8 | 152  (146 – 160) | 183  (176 – 192) | | 108  (104 – 114) | 130  (125 – 137) | |
| **Upper-middle** | 50.1 | 77.4  (74.6 – 79.6) | 136  (131 – 140) | | 56.2  (54.1 – 57.7) | 98.6  (95.0 – 101) | |
| **Lower-middle** | 29.8 | 133  (125 – 137) | 134  (126 – 139) | | 96.5  (90.6 – 99.9) | 97.3  (91.3 – 101) | |
| **Low** | 3.57 | 2.85  (2.72 – 2.97) | 125  (119 – 130) | | 2.13  (2.03 – 2.22) | 93.3  (88.8 – 96.9) | |
| Total life years gained are presented as medians with 95% credible intervals. Values are also presented as the median total life years gained per 1000 people vaccinated (first or second dose) with 95% credible intervals. Total undiscounted and discounted QALYs gained from averted COVID-19 deaths, infections and hospitalisations from COVID-19 vaccines are presented as medians with 95% credible intervals, with values also presented as total QALYs gained per 1000 people vaccinated (first or second dose) as medians with 95% credible intervals. Discounted QALYs were calculated by applying a 3% discount rate to the life years gained from averted COVID-19 deaths. Vaccine coverage is the proportion of the population with a full dose in the modelled countries by December 8, 2021. | | | | | | | |

Supplementary Table 3. Estimated infections and hospitalisations averted by COVID-19 vaccination

|  | **Vaccine coverage (%)** | **Infections averted** | | **Hospitalisations averted** | |
| --- | --- | --- | --- | --- | --- |
|  |  | **Total (billion)** | **Per 1000 people vaccinated** | **Total (million)** | **Per 1000 people vaccinated** |
| **Worldwide** | 38.8 | 1.50  (1.45 – 1.54) | 622  (602 – 638) | 51.5  (50.0 – 52.7) | 21.3  (0.0207 – 0.0218) |
| **World Bank income group** | | | | | |
| **High** | 68.8 | 0.562  (0.539 – 0.582) | 676  (648 – 700) | 21.4  (20.5 – 22.1) | 25.8  (24.7 – 26.6) |
| **Upper-middle** | 50.1 | 0.303  (0.295 – 0.314) | 532  (518 – 551) | 11.4  (11.0 – 11.7) | 20.0  (19.2 – 20.6) |
| **Lower-middle** | 29.8 | 0.619  (0.584 – 0.643) | 624  (589 – 648) | 18.4  (17.0 – 19.0) | 18.5  (17.1 – 19.1) |
| **Low** | 3.57 | 0.0198  (0.0190 – 0.0208) | 867  (833 – 909) | 0.472  (0.450 – 0.493) | 20.7  (19.7 – 21.6) |
| Total infections and hospitalisations averted from COVID-19 vaccines are presented as medians with 95% credible intervals, with values also presented as total infections or hospitalisations averted per 1000 people vaccinated (first or second dose) as medians with 95% credible intervals. Hospitalisations are considered as ‘potential’ hospitalisations, representing the incidence of averted severe cases that would have required hospitalisation; whether hospitalisation occurred or not is dependent on healthcare capacity. | | | | | |

**Supplementary Figure 1**. Global distribution of undiscounted monetized benefits from COVID-19 vaccination following extra-welfarist and welfarist approaches

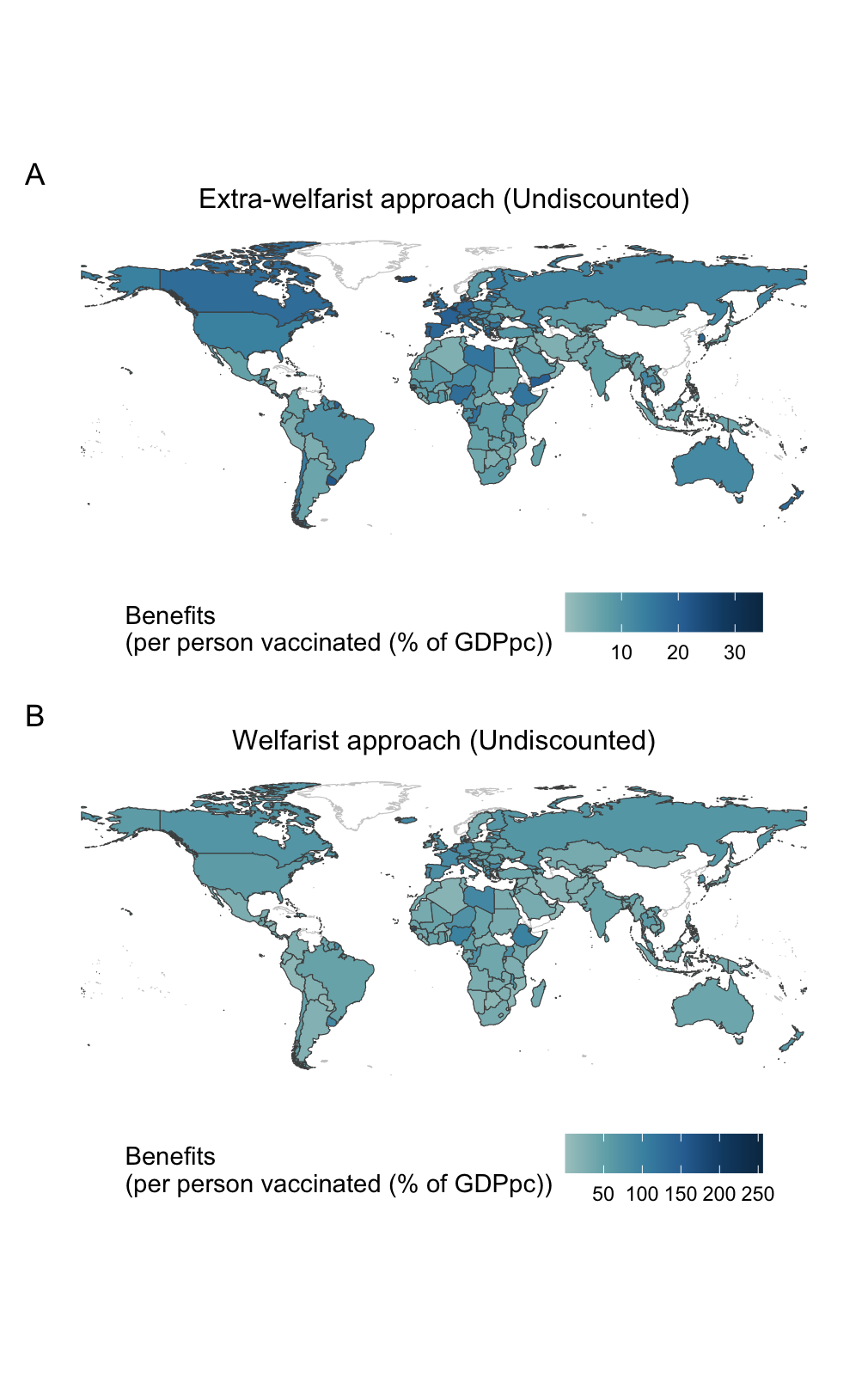

Figure 1A displays the sum of monetized benefits per person vaccinated under the extra-welfarist approach, including monetized QALYs gained, and productivity and healthcare costs averted. These results are expressed as a percentage of GDP per capita across different countries. Figure 1B displays the sum of the monetized benefits per person vaccinated under the welfarist approach, including VSLYs, expressed as a percentage of GDP per capita. Countries with no data available are shown outlined in light grey with no colour fill.

**Supplementary Figure 2.** Tornado plots displaying partial rank correlation coefficients (PRCCs) of key parameters included in the probabilistic sensitivity analysis for different outcomes. Higher absolute PRCCs indicate greater sensitivity of the outcome to the uncertainty in the corresponding parameter. The outcomes include: (A) discounted VSLYs, (B) discounted monetized QALYs, and (C) friction costs.

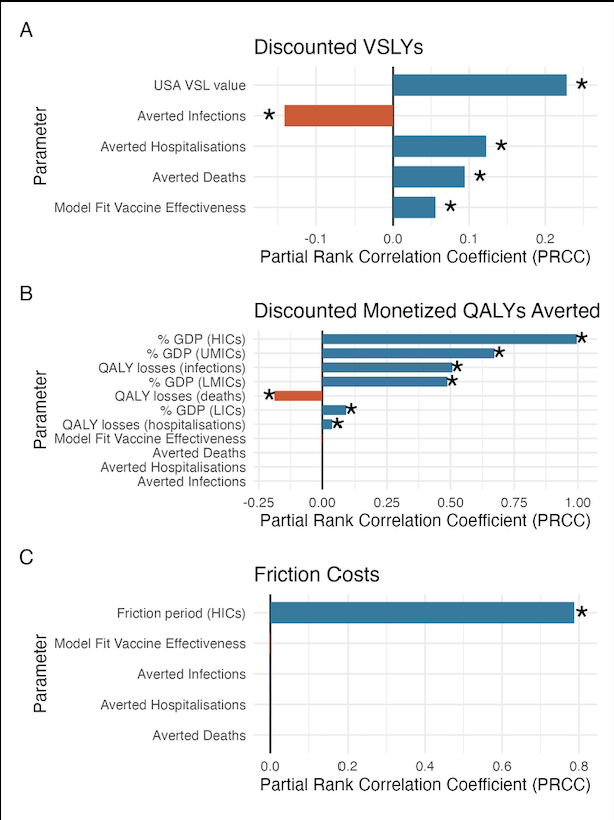

**Supplementary Figure 3.** Tornado plots displaying partial rank correlation coefficients (PRCCs) of key parameters included in the probabilistic sensitivity analysis for different outcomes. Higher absolute PRCCs indicate greater sensitivity of the outcome to the uncertainty in the corresponding parameter. The outcomes include: (A) undiscounted VSLYs, and (B) undiscounted monetized QALYs.

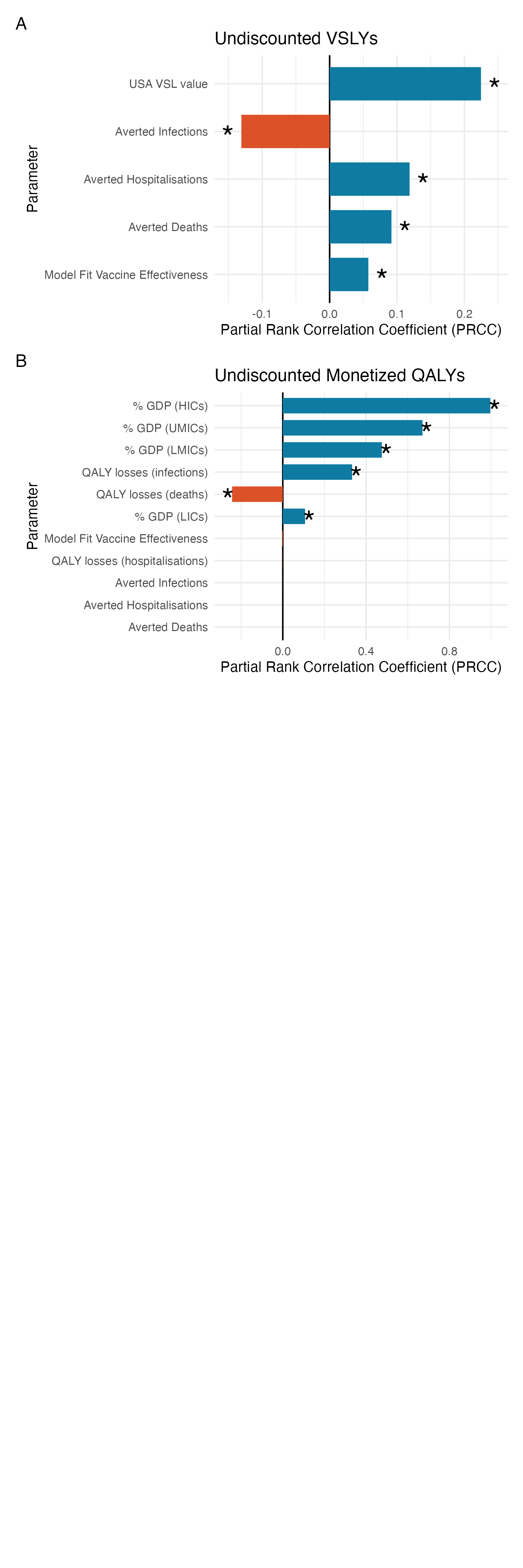

20.	Florio M, Gamba S, Pancotti C. Mapping of long-term public and private investments in the development of COVID-19 vaccines.
